# Association of echocardiographic cardiac valve calcification with major adverse cardiovascular events in patients on acute myocardial infarction

**DOI:** 10.1101/2024.10.15.24315566

**Authors:** Chuyun Chen, Na Wang, Yang Yu, Jia Jia, Fangfang Fan, Ying Yang, Yan Zhang

**Author notes:** Correspondence to: Yan Zhang, Department of Cardiology, Peking University First Hospital, No. 8 Xishiku Street, Xicheng District, Beijing 100034, China.

## Abstract

**BACKGROUND:** The role of cardiac valve calcification (CVC) in secondary prevention post-acute myocardial infarction (AMI) remains unclear. This study investigated the association between CVC and major adverse cardiovascular events (MACE) in AMI.

**METHODS AND RESULTS:** AMI patients hospitalized in Peking University First Hospital were consecutively enrolled. Participants were grouped according to the measurement of transthoracic echocardiography on admission. According to aortic valve calcification (AVC) and mitral annular calcification (MAC), patients were divided into Group 1 (with no valve calcification), Group 2 (with single valve calcification) and Group 3 (with both valve calcification). The primary endpoint was MACE (defined as a composite of nonfatal stroke, nonfatal myocardial infarction and cardiac death). A cohort of 911 AMI patients was recruited in this retrospective study. There were 513(56.31%), 337(36.99%) and 61(6.70%) in Group 1, Group 2 and Group 3, respectively. During follow-up (median, 5.11 years), 277(30.41%) patients developed MACE, 94(10.32%) had nonfatal stroke, 155(17.01%) had recurrent myocardial infarction (MI) and 106(11.64%) were identified as cardiac death. Comparing with Group 1, the adjusted hazard ratios (HR) for primary endpoint and recurrent MI in Group 3 was 1.61 (1.06, 2.43) and 1.70 (1.00, 2.89), respectively. And the adjusted HR of primary endpoint and cardiac death in moderate and severe AVC group was 1.50 (1.01, 2.23) and 1.97 (1.10, 3.51) compared with non-AVC group.

**CONCLUSIONS:** AMI patients with CVC were at significantly increased risk of MACE, although existing differences of the position and the severity of calcification. Screening for AVC and MAC is necessary in secondary prevention.

**RESEARCH PERSPECTIVE:** 

**What Is New?:**

- This study is the first to detail the association between cardiac valve calcification (CVC), specifically aortic valve calcification (AVC) and mitral annular calcification (MAC), with major adverse cardiovascular events (MACE) in patients after an acute myocardial infarction (AMI).
- It also identifies the concurrent presence of MAC and AVC as an independent predictor of MACE.

**What Are the Clinical Implications?:**

- The stud y suggests that screening for AVC and MAC post-AMI could help identify patients at higher risk of MACE, leading to more targeted preventive care and potentially improving outcomes.

---

Aortic valve calcification (AVC), also known as calcific aortic valve disease (CAVD) is a prevalent condition among the elderly population. Between 1990 and 2019, there has been a significant escalation in the global incidence, prevalence, and mortality rates of CAVD, increasing by 3.51-fold, 4.43-fold, and 1.38-fold, respectively ^1^. A Chinese study reveals that in 2010, approximately 25,621,503 individuals were affected by valvular heart disease, with a weighted prevalence of 3.8% ^2^. Mitral annulus calcification (MAC), also a subtype of valvular heart disease, is frequently linked to degenerative alterations. A comprehensive global epidemiological study indicates that 24 million individuals were afflicted with degenerative mitral valve disease ^3^.

Valvular calcification is a prevalent condition among patients undergoing maintenance dialysis for end-stage renal disease, often attributed to dysregulation in calcium and phosphorus metabolism. Studies indicate that the detection rates for AVC and MAC in this patient population is 55%-69% and 40%-60%, respectively ^4, 5^. The prognostic significance of cardiac valve calcification in predicting major adverse cardiovascular events (MACE) has been extensively studied, particularly in the context of maintenance dialysis patients with end-stage renal disease. A meta-analysis included 22 chronic kidney disease (CKD) researches shows that CVC (the presence of both MAC and AVC) increase the risk of all-cause and cardiovascular mortality in CKD patients. CVC is slightly associated with cardiovascular events in CKD patients (HR: 2.1, 95% CI: 0.96–4.59) ^6^.

CVC shares pathological characteristics and risk factors with atherosclerosis, leading to its consideration as a marker for subclinical atherosclerosis and an independent predictor of adverse outcomes in primary prevention ^7^. However, research on the predictive value of CVC in the secondary prevention setting, particularly in patients with acute myocardial infarction (AMI), remains limited. The objective of this study is to explore the correlation between valve calcification and MACE in AMI.

## METHODS

### Study Population

From January 2010 to December 2018, patients admitted for AMI in the Department of Cardiology of Peking University First Hospital were recruited. The inclusion criteria were: (1) age ≥18 years; (2) diagnosed for AMI; (3) with complete clinical data in electronic medical record and biological sample; (4) echocardiographic images storaged after admission that allowed perfect measurement of AVC and MAC. The exclusion criteria were as follows: (1) less than 18 years; (2) with incomplete clinical data or no biological sample; (3) angina; (4) with unsatisfied echocardiographic images. The study protocol was approved by the Peking University First Hospital ethics committee and conducted in accordance with the Declaration of Helsinki. Written informed consent was required for inclusion in the study.

Demographic and clinic characteristics of all patients included age, sex, alcohol consumption, disease (i.e., transient ischemic attacks, ischemic stroke, or hemorrhagic stroke), type of myocardial infarction, percutaneous coronary intervention (PCI), and use of aspirin, clopidogrel or ticagrelor, beta-blockers, angiotensin-converting enzyme inhibitors/angiotensin receptor antagonists and statins. Furthermore, laboratory measurements included total cholesterol, low-density lipoprotein cholesterol (LDL-C), high-density lipoprotein cholesterol (HDL-C), triglycerides and creatinine. All the information above were extracted from the electronic health records.

### Echocardiography

Echocardiography were measured in all AMI patients on admission using an ultrasound system (Vivid-7; General Electric, Boston, MA, USA) with a 3-MHz transducer and digitally stored. The presence and severity of AVC and MAC was detected and recorded by two experienced physicians who were blinded to the clinical data of the patients. The severity of AVC was classified into three levels: (1) with no calcification; (2) mild calcification (small isolated spots); (3) moderate calcification (multiple larger spots), or severe calcification (extensive thickening and calcification of all cusps) ^8^. Three levels were also applied in the classification of MAC: (1) with no calcification; (2) mild calcification (involving one-third or less of the annulus); (3) moderate calcification (involving one-third to one-second of the annulus) or severe calcification (involving more than one-second of the annulus) ^9^. Moreover, all the subjects were divided into three groups: Group 1 (with no valve calcification), Group 2 (with single valve calcification) and Group 3 (with both valve calcification). The evaluation of AVC and MAC were re-measured in 15 randomly selected patients by the same observer who was blinded to their previous measurements with a one-week interval between measurements to assess intraobserver reliability and by a second observer blinded to the measurements obtained by the first observer to assess inter-observer reliability. Inter-observer and intra-observer agreement was evaluated using the kappa statistic (poor, κ≤0.40; moderate, κ=0.40–0.60; good, κ=0.60–0.80; and excellent, κ>0.80). Inter-observer agreement was excellent for both AVC (κ=0.887) and MAC (κ=0.842), as was intra-observer agreement (κ=0.825 and κ=0.815, respectively) (all P<0.05).

### Outcomes

The primary endpoint was MACE (defined as a composite of nonfatal stroke, nonfatal myocardial infarction and cardiac death). And the secondary endpoint included stroke, myocardial infarction and cardiac death. The outcome data originated from the Chinese Center for Disease Control and Prevention and the Beijing Municipal Health Commission. Information were also recorded by telephone interviews with patients or their family members performed by trained reviewers who were blinded to the echocardiography measurements. All the patients were followed-up (with no missing) after initial echocardiography until December 31, 2021 or the occurrence of endpoint of this study.

### Statistical Analysis

The variables are expressed as the mean ± standard deviation, median (interquartile range), or number (percentage). Student’s *t*-test, chi-squared test, or Fisher’s exact test were used appropriately to make comparisons among the groups. Patients with moderate and severe calcification were integrated in consideration of the small quantity of the patients in the severe AVC and severe MAC groups. Time-to-event data were estimated using the Kaplan-Meier method with application of the log-rank test to make comparisons. The effect of AVC, MAC and CVC on the occurrence of endpoints were evaluated by the univariable and mutivariable Cox proportional risk regression models, with hazard ratio (HR) and 95% confidence interval (CI) turning out. Factors with a P-value of <0.1 in univariate Cox regression and those that were clinically considered meaningful were selected for further analysis in multivariate Cox proportional hazard models. Subgroup analysis was used to evaluate the heterogeneity of exposure effect on the endpoint. The prespecified subgroups were as follows: age (<65 or ≥65 years), sex, smoking, hypertension, diabetes mellitus, PCI, type of AMI, and CKD. All statistical analyses were performed using R software (version 4.3.3, http://www.R-project.org) and Empower (www.empowerstats.com, X&Y Solutions, Inc, Boston, MA, USA). A two-tailed P-value of <0.05 was considered statistically significant.

## RESULTS

### Baseline Characteristics

As shown in **Table 1**, a total of 911 AMI patients were included. The mean age was 63.02±12.44 years and 217(23.82%) were women. After echocardiographic assessments, 533(58.51%) patients were without AVC, 299 patients (32.82%) were found to have mild AVC, and 79(8.67%) for moderate AVC and severe AVC; 830(91.11%) were without MAC, 51(5.60%) patients were found to have mild MAC, and 30(3.29%) for moderate and severe MAC. There were 513(56.31%), 337(36.99%) and 61(6.70%) in Group 1, Group 2 and Group 3, respectively. During follow-up (median, 5.11 years), 277(30.41%) cases of MACE occurred, 94(10.32%) had nonfatal stroke, 155(17.01%) had recurrent myocardial infarction (MI) and 106(11.64%) were identified as cardiac death. Patients in Group 3 were older and were more likely to be women. They also tended to have comorbidities such as hypertension, diabetes, cerebrovascular disease and chronic kidney disease (**Table 1**). The patient characteristics were shown stratified by the severity of AVC and MAC or MACE in **Supplemental Table S1, Supplemental Table S2** and **Supplemental Table S3**.

**Table 1.**
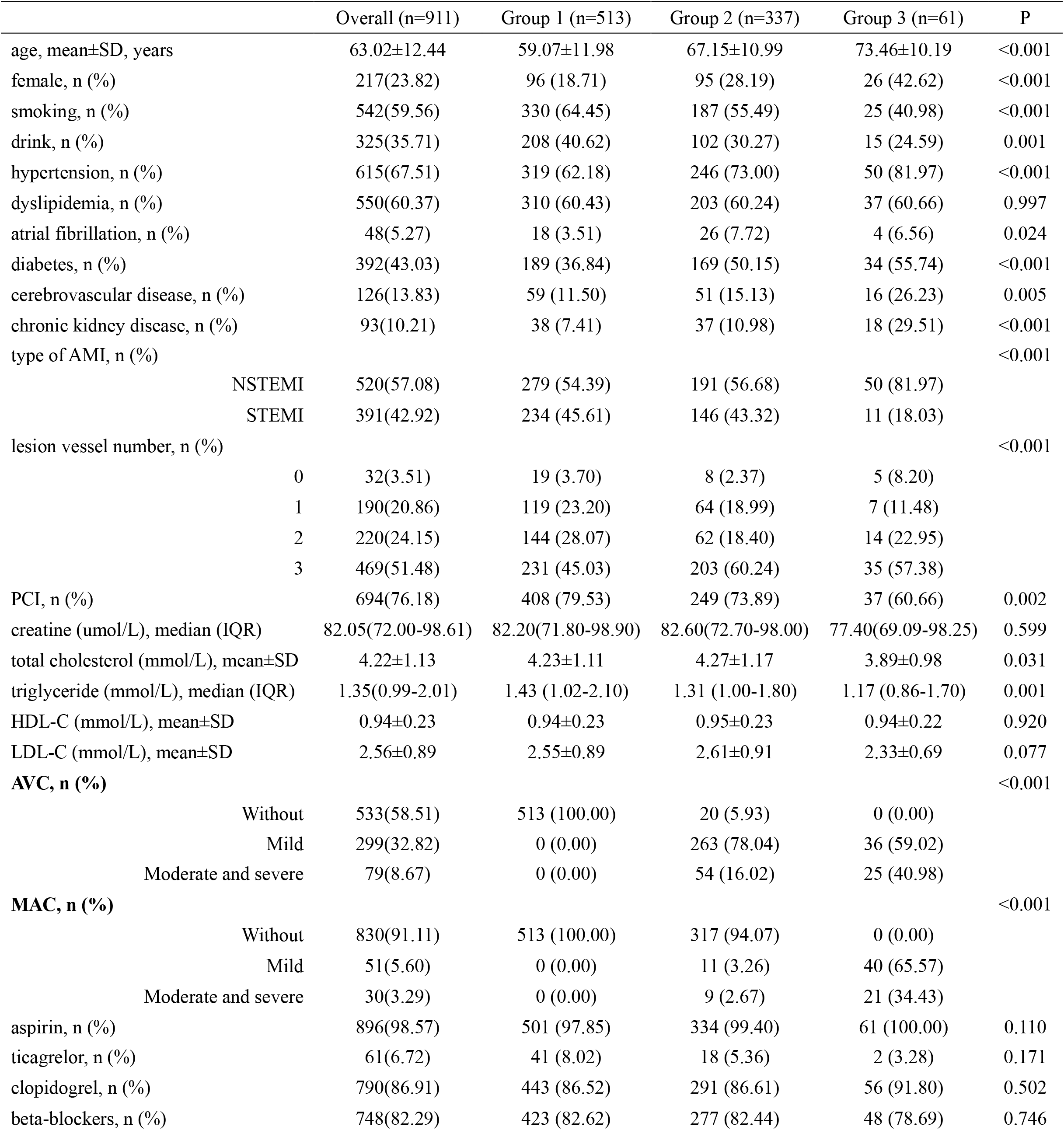
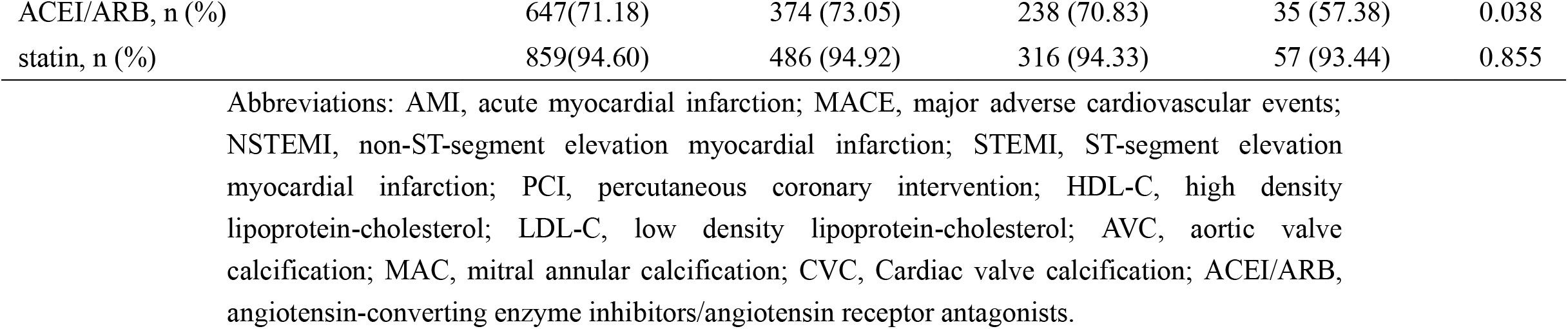
Baseline clinical characteristics of all the AMI patients

### Primary endpoint

During follow-up (median, 5.11 years), 277(30.41%) cases of MACE occurred, 94(10.32%) had nonfatal stroke, 155(17.01%) had recurrent myocardial infarction (MI) and 106(11.64%) were identified as cardiac death. The cumulative risk curves of MACE are presented in **Figure 1, Figure 2** and **Figure 3**. Patients with moderate and severe AVC were more likely to have MACE (adjusted HR=1.50, 95% CI: 1.01-2.23, P=0.045), comparing with non-AVC group (**Table 2**). Patients in Group 3 were more likely to have MACE (adjusted HR=1.61, 95% CI: 1.61-2.43, P=0.025), comparing with Group 1 (**Table 4**). And these above differences were not the case for moderate and severe MAC (adjusted HR=1.54, 95% CI: 0.89-2.65, P=0.125) (**Table 3**).

**Table 2.**
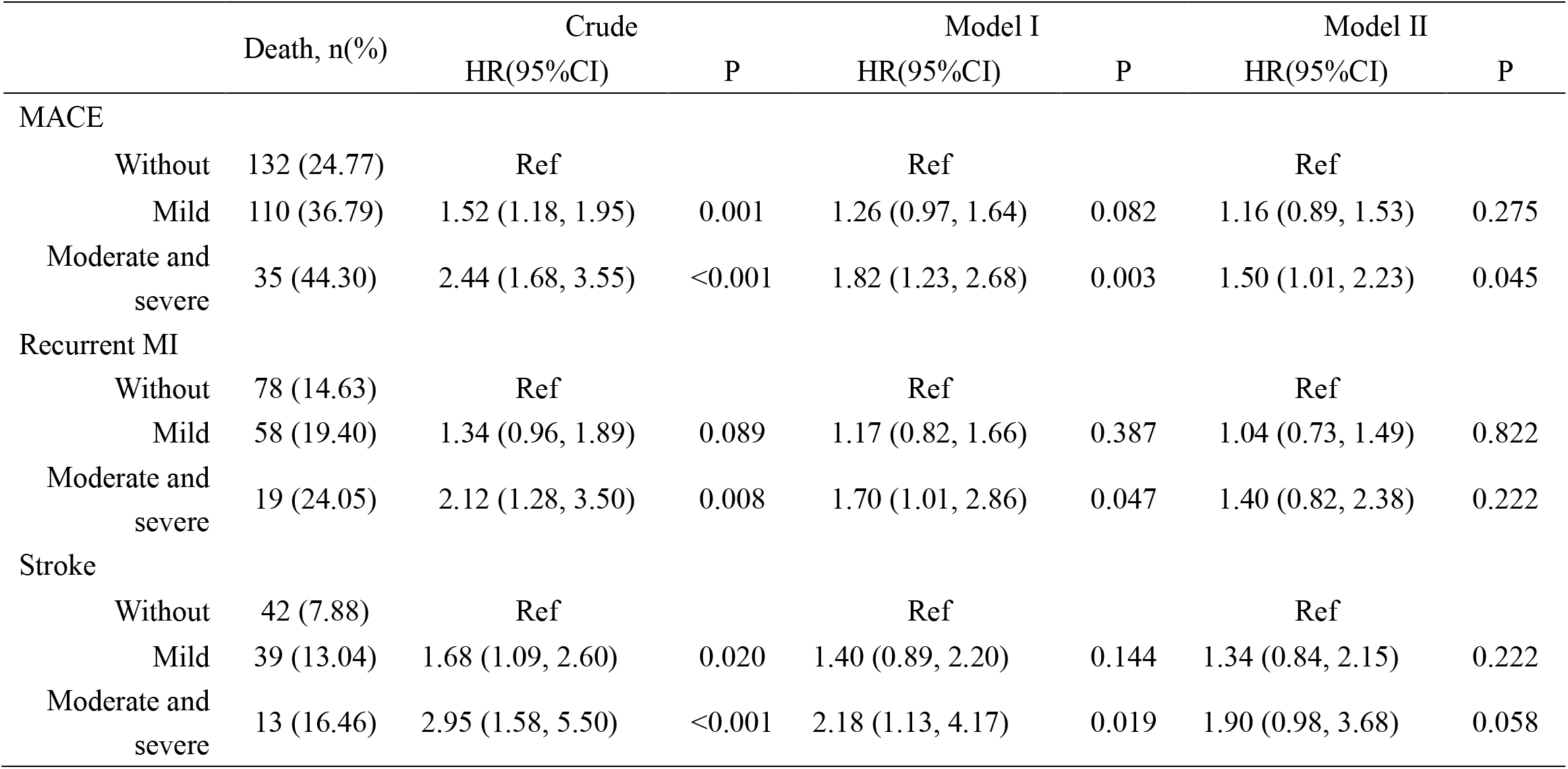
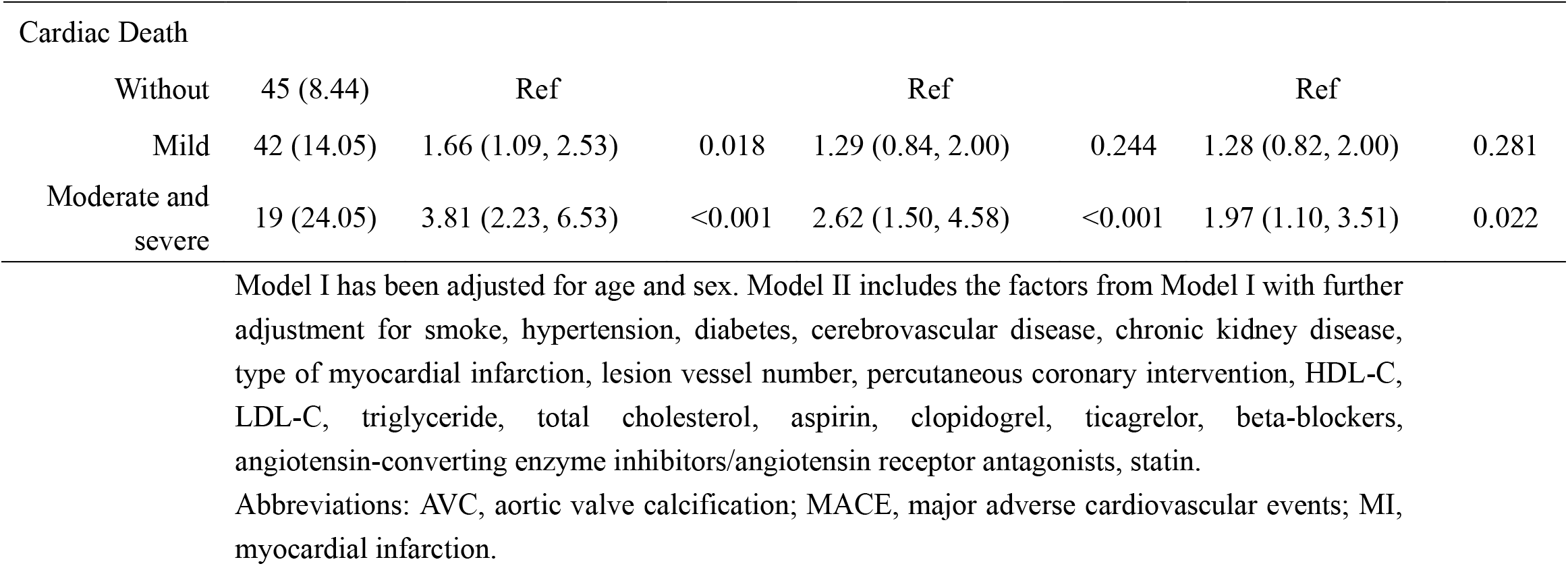
The association between AVC and MACE.

**Table 3.**
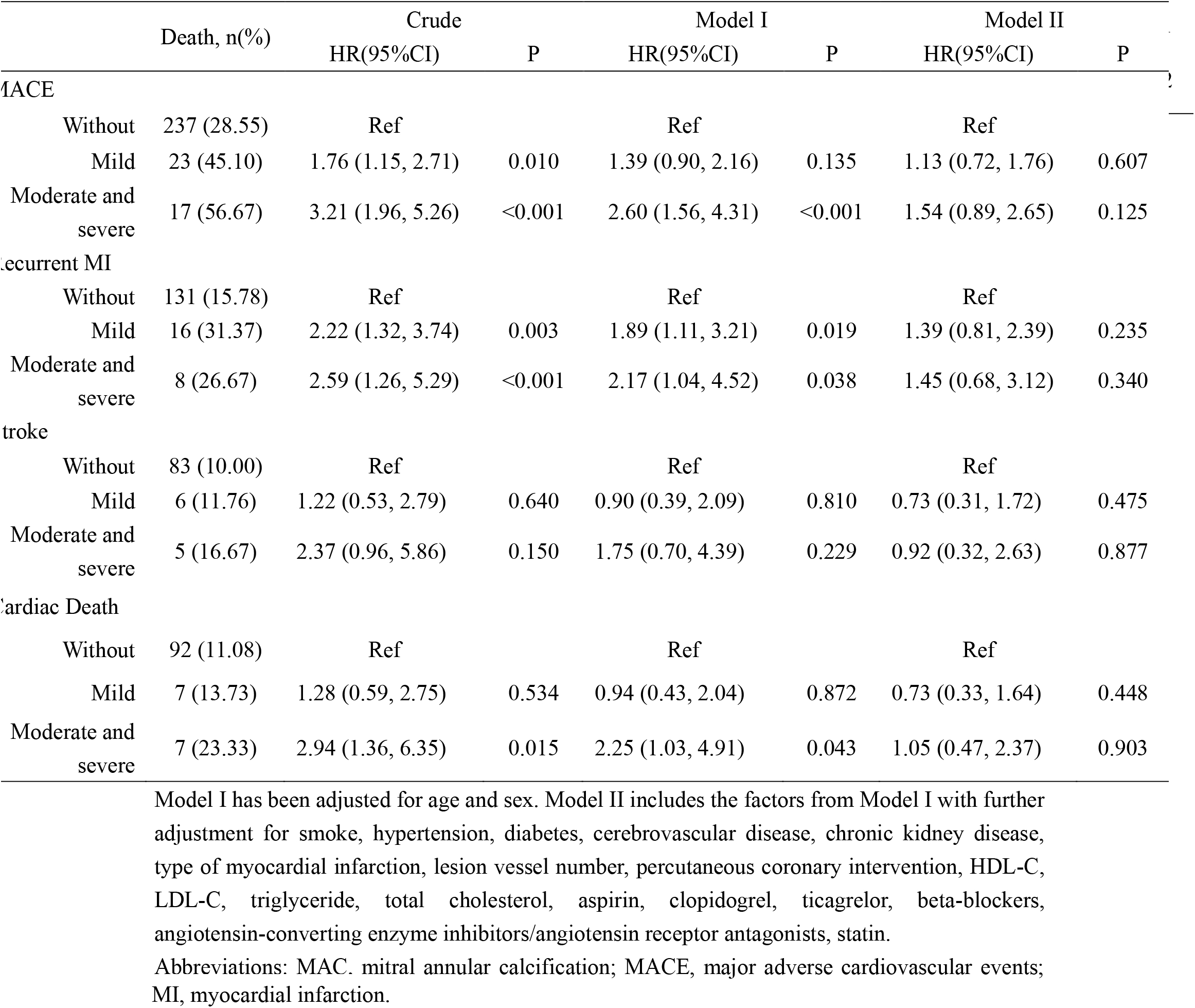
The association between MAC and MACE.

**Table 4.** The association between CVC and MACE.

|  | Death, n(%) | Crude<br>HR(95%CI) | P | Model I<br>HR(95%CI) | P | Model II<br>HR(95%CI) | P |
| --- | --- | --- | --- | --- | --- | --- | --- |
| **MACE** |  |  |  |  |  |  |  |
| Group 1 | 127 (24.76) | Ref |  | Ref |  | Ref |  |
| Group 2 | 115 (34.12) | 1.41 (1.10, 1.82) | 0.007 | 1.18 (0.91, 1.54) | 0.208 | 1.08 (0.82, 1.42) | 0.587 |
| Group 3 | 35 (57.38) | 3.28 (2.26, 4.78) | <0.001 | 2.34 (1.57, 3.47) | <0.001 | 1.61 (1.06, 2.43) | 0.025 |
| **Recurrent MI** |  |  |  |  |  |  |  |
| Group 1 | 75(14.62) | Ref |  | Ref |  | Ref |  |
| Group 2 | 59(17.51) | 1.22 (0.87, 1.71) | 0.258 | 1.07 (0.75, 1.52) | 0.708 | 0.93 (0.65, 1.34) | 0.694 |
| Group 3 | 21(34.43) | 3.20 (1.97, 5.21) | <0.001 | 2.50 (1.50, 4.19) | <0.001 | 1.70 (1.00, 2.89) | 0.050 |
| **Stroke** |  |  |  |  |  |  |  |
| Group 1 | 41 (7.99) | Ref |  | Ref |  | Ref |  |
| Group 2 | 43 (12.76) | 1.66 (1.08, 2.55) | 0.020 | 1.37 (0.88, 2.14) | 0.165 | 1.33 (0.84, 2.11) | 0.226 |
| Group 3 | 10 (16.39) | 2.59 (1.30, 5.17) | 0.006 | 1.76 (0.85, 3.65) | 0.126 | 1.19 (0.55, 2.56) | 0.662 |
| **Cardiac Death** |  |  |  |  |  |  |  |
| Group 1 | 43 (8.38) | Ref |  | Ref |  | Ref |  |
| Group 2 | 51 (15.13) | 1.84 (1.23, 2.76) | 0.003 | 1.44 (0.95, 2.19) | 0.089 | 1.34 (0.87, 2.07) | 0.184 |
| Group 3 | 12 (19.67) | 2.89 (1.52, 5.48) | <0.001 | 1.83 (0.94, 3.57) | 0.074 | 1.21 (0.61, 2.40) | 0.576 |

Model I has been adjusted for age and sex. Model II includes the factors from Model I with further adjustment for smoke, hypertension, diabetes, cerebrovascular disease, chronic kidney disease, type of myocardial infarction, lesion vessel number, percutaneous coronary intervention, HDL-C, LDL-C, triglyceride, total cholesterol, aspirin, clopidogrel, ticagrelor, beta-blockers, angiotensin-converting enzyme inhibitors/angiotensin receptor antagonists, statin.
Abbreviations: CVC, Cardiac valve calcification; MACE, major adverse cardiovascular events; MI, myocardial infarction.

**Fig. 1.**
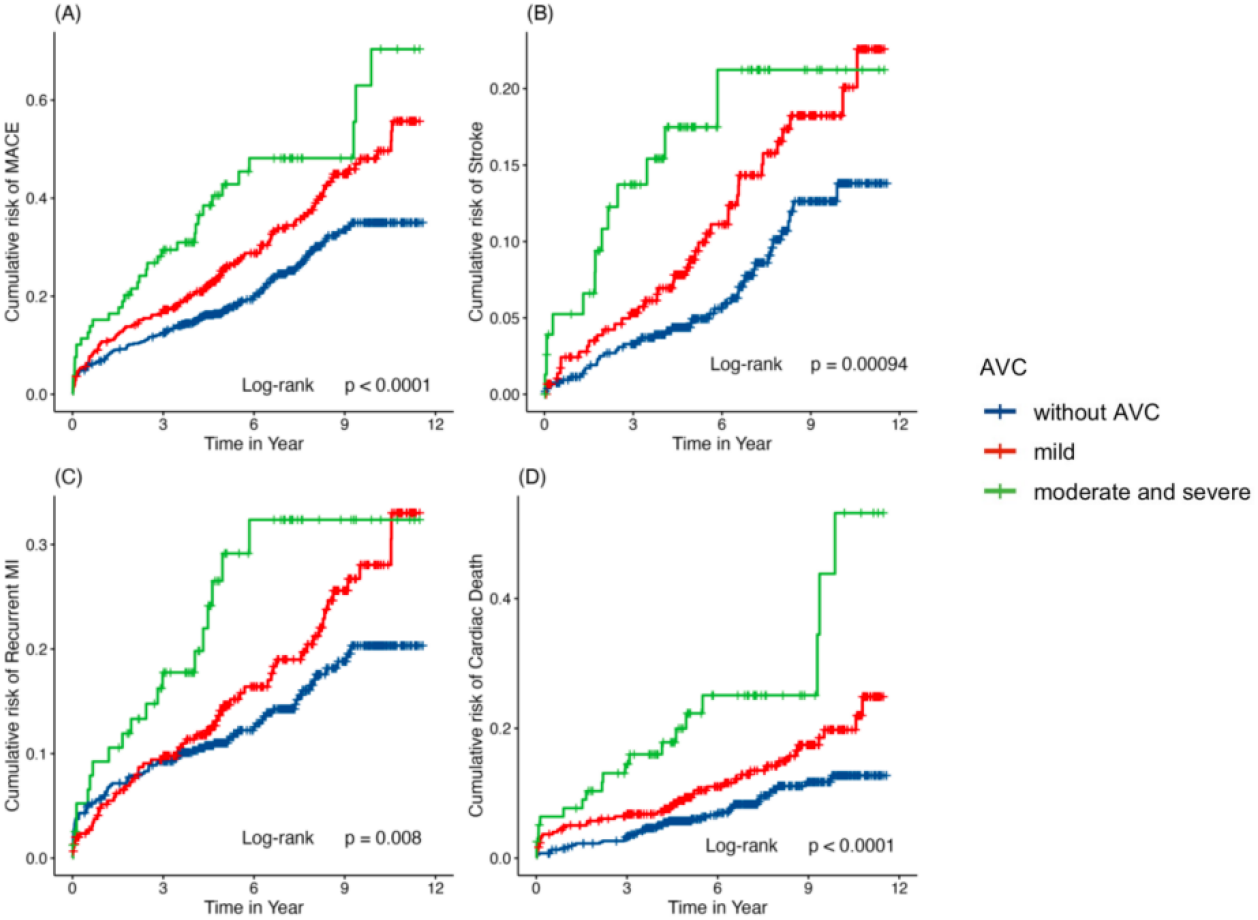
The cumulative risk curve of AVC for MACE (A), Stroke (B), Recurrent MI (C) and Cardiac Death (D). Abbreviations: AVC, aortic valve calcification; MACE, major adverse cardiovascular events; MI, myocardial infarction.

**Fig. 2.**
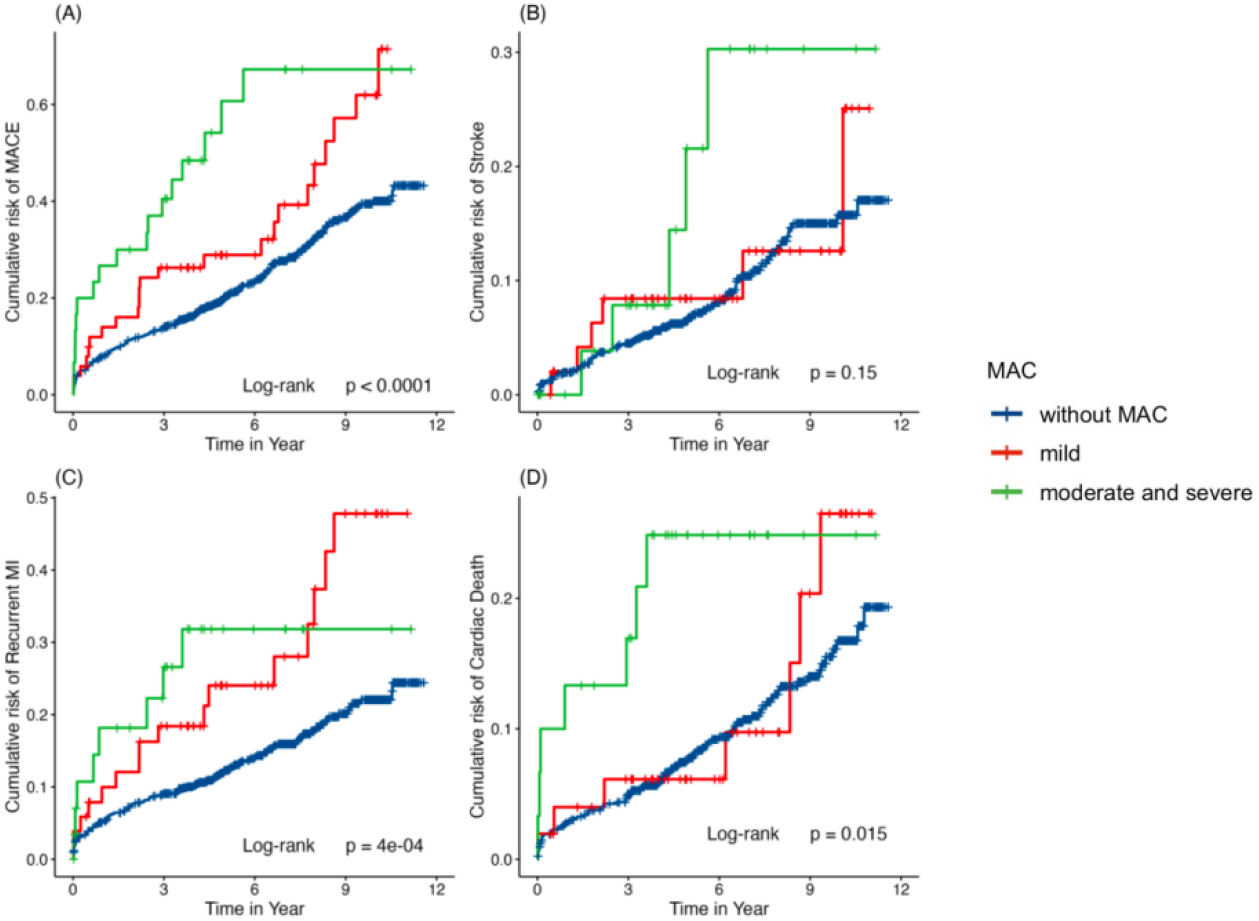
The cumulative risk curve of MVC for MACE (A), Stroke (B), Recurrent MI (C) and Cardiac Death (D). Abbreviations: MVC, aortic valve calcification; MACE, major adverse cardiovascular events; MI, myocardial infarction.

**Fig. 3.**
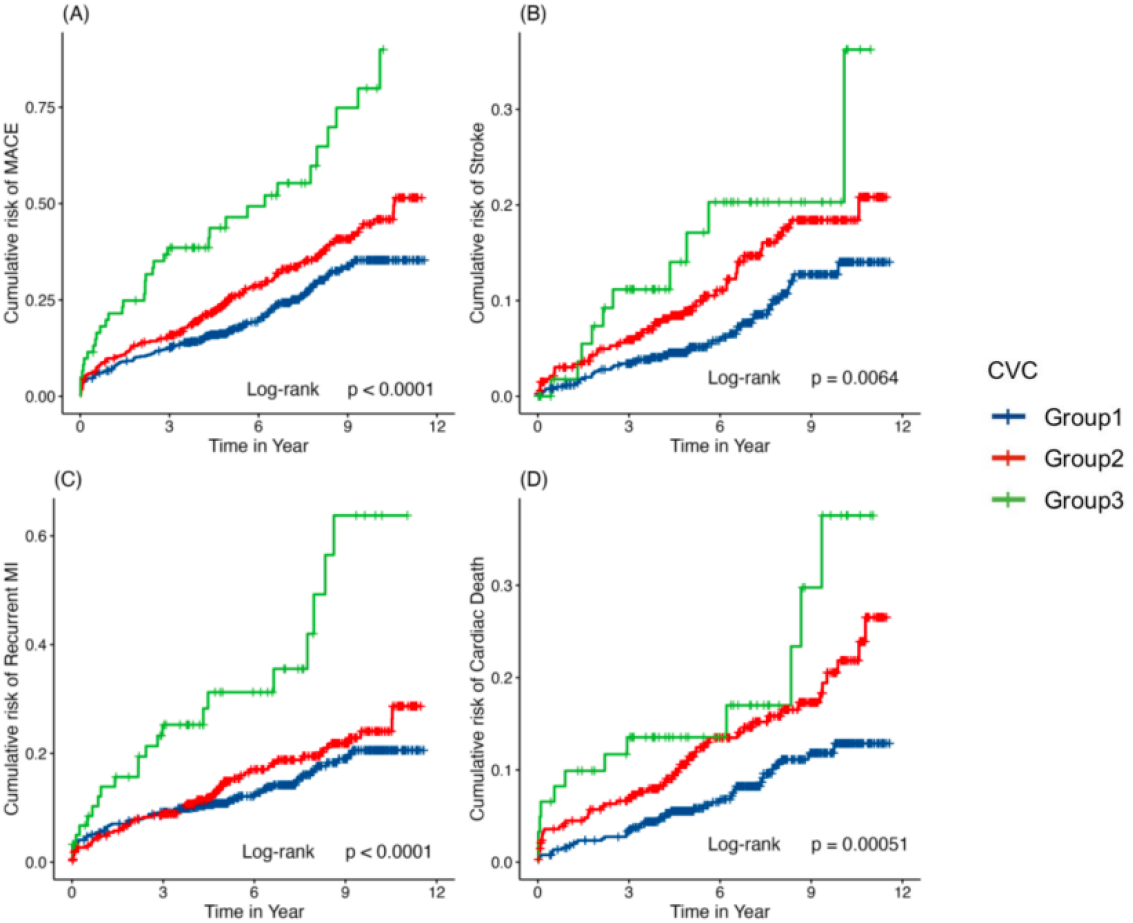
The cumulative risk curve of cardiac valve calcification for MACE (A), Stroke (B), Recurrent MI (C) and Cardiac Death (D). Abbreviations: MACE, major adverse cardiovascular events; MI, myocardial infarction.

### Subgroup analysis

The association between AVC and MACE was consistent across the prespecified subgroups (**Supplemental Table S4**). The association between CVC and MACE was further consistent across the prespecified subgroups except for hypertension (**Supplemental Table S5**). Hypertension did not affect the overall results, but the association between CVC and MACE seemed to be more strong in patients without hypertension.

### Secondary endpoint

As shown in **Table 2**, patients with moderate and severe AVC were more likely to occur cardiac death compared with patients with no AVC (adjusted HR=1.97, 95% CI: 1.10-3.51, P=0.022). The incidence of recurrent myocardial infarction or stroke was not associated with the presence of moderate and severe AVC. Moreover, of notes were the correlation between Group 3 and recurrent myocardial infarction (adjusted HR=1.70, 95% CI: 1.00-2.89, P=0.050) (**Table 4**). However, similar relationship did not present in moderate and severe MAC (**Table 3**).

## DISCUSSION

This retrospective cohort study investigated the correlation between valve calcification and MACE in AMI patients under the circumstance of secondary prevention. The study found that moderate or severe AVC was significantly associated with MACE (HR= 1.50, 95%CI: 1.01-2.23, P= 0.045). Moreover, the concurrent presence of MAC and AVC was identified as an independent predictor of MACE (HR= 1.61, 95%CI: 1.06-2.43, P= 0.025) in AMI.

CVC was recognized as a prognostic indicator of adverse prognosis in CKD ^6, 10^. In dialysis patients, vascular calcifications, particularly AVC, emerged as a non-traditional cardiovascular risk factor and a marker of cardiovascular disease progression. Research utilizing transthoracic echocardiography to identify AVC in CKD consistently demonstrated association between vascular calcifications and cardiovascular events ^11-13^.

Nevertheless, the prognostic significance of CVC in AMI was less extensively discussed in comparison to CKD. CVC shared similar pathophysiological characteristics with atherosclerosis, indicating a potential role in the development of cardiovascular complications ^14, 15^. Evidence based on the Multi-Ethnic Study of Atherosclerosis (MESA) study revealed a synergistic interaction between valve calcification and coronary calcification in the development of cardiovascular diseases. This underscored the importance of managing and detecting AVC in clinical practice, which could be instrumental in preventing adverse outcomes ^16^. An American study involving veterans with moderate-to-high atherosclerotic cardiovascular risk demonstrated the predictive value of AVC, identified by computed tomography (CT), for mortality, nonfatal MI, and nonfatal cerebrovascular accident, independent of coronary calcification or severe aortic valve stenosis ^17^. Two Japanese studies, focusing on patients with suspected coronary heart disease or coronary artery disease, also observed similar correlations ^18, 19^. However, in the context of secondary prevention, direct clinical evidence regarding valve calcification remained limited. Ge’s team indicated that AVC was beneficial for risk stratification in AMI patients post-percutaneous coronary intervention (PCI). The presence of AVC significantly increased the risk of MACE (HR=1.44, 95%CI: 1.19-1.75, P < 0.001) ^20^. Another study by his team showed that AVC was also an independent predictor for periprocedural myocardial injury, defined as hs-cTnT after coronary intervention higher than 99th percentile upper reference limit ^21^. Although the two aforementioned research were conducted in China, we provided a more detailed classification of AVC severity (none, mild, moderate, and severe) and discovered that the moderate and severe AVC groups were at a higher risk. Furthermore, we considered the prevalence of both AVC and MAC simultaneously, examining the number of valve calcification on outcomes.

The impact of mitral annular calcification (MAC) on adverse events remained a subject of debate. A cross-sectional study involving end-stage renal disease patients, which considered cardiovascular disease and traditional risk factors, reported negative findings ^22^. However, a large-scale cohort study from the Mayo Clinic identified a higher mortality risk in patients with MAC, with the risk further increasing in the presence of both MAC and mitral valve dysfunction ^23^. While the proportion of patients with MAC in our study was only 8.89%, which was lower than that in the previous studies ^23-25^. Additionally, several studies reported sex differences, with the correlation between CVC and poor prognosis appearing to be stronger in women ^26^. Distinct biomarker signatures for valve calcification have been observed in different sexes, with men showing oxidative stress and women exhibiting both oxidative stress and inflammation. These factors might influence cardiovascular complications or adverse events ^27^. The lower proportion of women (23.82%) in our study could partially explain the observed negative relationship between MAC and MACE to some extent. Further investigation was needed to elucidate the role and mechanisms of MAC in cardiovascular outcomes. It was noteworthy that while most studies defined CVC as the presence of at least one calcified valve ^13, 28, 29^, our study provided a more detailed classification (three groups), which aided in distinguishing the roles of aortic valve calcification (AVC) and MAC in secondary prevention.

Additionally, the potential link between stroke and valve calcification was examined in previous studies, yielding mixed results. The Rotterdam Study, which focused on community-dwelling elderly individuals with a mean age of 69.6 years, indicated that AVC was not significantly associated with an increased risk of stroke ^30^. Similarly, no significant correlation was observed between AVC and MAC in relation to stroke in patients undergoing coronary artery bypass grafting ^25^. However, a separate cardiovascular health study utilizing magnetic resonance imaging revealed a direct relationship between the severity of valvular calcification and the presence of brain infarcts ^31^. In summary, the connection between valve calcification and stroke risk remains equivocal. There was a need for more extensive, prospective studies to clarify this relationship in the future ^32^.

This study has some unavoidable limitations. First, the nature of observational cohort design meant unsure cause-effect relationship. Second, the relatively low proportion of female patients, at 23.82%, may limit the generalizability of the findings.

## CONCLUSIONS

The risk for major adverse cardiovascular events after AMI is higher in the presence of both AVC and MAC. And patients with moderate and severe AVC tend to experience poor prognosis. Screening for CVC may provide additional clinical value on AMI in secondary prevention.

## Data Availability

The data that support the findings of this study are available from the corresponding author upon reasonable request after the request is submitted and formally reviewed and approved by the ethics committee of Peking University First Hospital. Requests to access the datasets should be directed to.

## ARTICLE INFORMATION

### Affiliations

Department of Cardiology (C.C., N.W., Ya.Y., J.J., F.F., Yi.Y., Y.Z.) and Institute of Cardiovascular Disease (J.J., F.F., Y.Z.), Peking University First Hospital, Beijing, China.

## Acknowledgments

None

## Sources of Funding

This study was financed by the National Key Research and Development Program of China (2021YFC2500503).

### Disclosures

None.

### Supplementary Information

Tables S1–S5

